# Association of age-related spontaneous internal jugular vein reflux with cognitive impairment and incident dementia

**DOI:** 10.1101/2023.06.21.23291726

**Authors:** Utako Adachi, Sono Toi, Megumi Hosoya, Takao Hoshino, Misa Seki, Hiroshi Yoshizawa, Yukiko Tsutsumi, Kenji Maruyama, Kazuo Kitagawa

**Author notes:** Correspondence to: Kazuo Kitagawa, MD Department of Neurology, Tokyo Women’s Medical University Hospital 8-1, Kawada-cho, Shinjuku-ku, 162-8666 Tokyo, Japan.

## Abstract

**Background and purpose:** The vascular aspect of cognitive impairment focuses on cerebral small-vessel disease and arterial circulation. However, it remains unclear whether changes in the venous circulation contribute to cognitive decline. This study aimed to clarify whether the spontaneous jugular vein reflux (JVR) is associated with cognitive impairment and incident dementia.

**Methods:** The Tokyo Women’s Medical University Cerebrovascular Disease registry is a prospective observational registry in which patients with any evidence of cerebral vessel disease on magnetic resonance imaging (MRI) were consecutively enrolled between October 2015 to July 2019. We employed carotid duplex sonography to measure the internal jugular vein (IJV). The subjects were classified into two groups based on the degree of JVR on either side: none, mild (JVR(-) group) and moderate, severe (JVR (+) group) JVR. They underwent both the Mini-mental State Examination (MMSE) and Montreal Cognitive Assessment-Japanese (MoCA-J) global tests. Their cognitive status was prospectively assessed until March 2022.

**Results:** Among 906 patients with an MMSE score ≥ 24 enrolled in the registry, 302 underwent duplex sonography of the IJV. Among them, 91 had spontaneous JVR on either side. Both MMSE and MoCA-J were significantly lower in patients with JVR (+) group than in the JVR (-) group. After the adjustment for age, sex, years of education, risk factors, and MRI findings, including small-vessel disease and medial temporal atrophy, intergroup differences in MoCA-J remained significant. Among the cognitive subdomains, median executive function and memory scores were significantly lower in the JVR (+) group than in the JVR (-) group. During the median 5.2-year follow-up, 11 patients with incident dementia (Alzheimer disease: 10, vascular dementia: 1) were diagnosed. Patients with severe JVR were significantly more likely to be diagnosed with dementia (log-rank test, P= 0.031).

**Conclusions:** Spontaneous IJV reflux especially severe JVR, was associated with global cognitive function, and potentially with incident dementia.

**Clinical Trial Registration:** URL:https://www.umin.ac.jp. Unique identifier: UMIN000026671.

## Introduction

The vascular aspect on cognitive impairment and dementia has attracted considerable attention because of the possibility of preventing dementia^1^. Aging influences vascular function in the arterial and venous systems. Although previous studies mostly focused on cerebral small-vessel disease (SVD) and arterial circulation in cognitive impairment, few examined the involvement of age-related venous dysfunction in cognitive function. The cerebral venous system plays an important role in normal brain function^2,3^, and an increase in cerebral vein pressure due to arterial venous fistula^4^ or cerebral venous thrombosis^5^ contributes to white matter lesions, cognitive decline and dementia. The internal jugular veins (IJVs) are the main drainage route for cerebral venous outflow^6,7^, and its hemodynamics can be examined using a non-invasive ultrasound method^8^. The flow of the IJVs usually moves from the brain to the heart and anterograde; however, the frequency of the retrograde flow pattern, jugular vein reflux (JVR), increases with aging^8^. Although findings in multiple sclerosis are controversial^9^, the association of JVR with normal-pressure hydrocephalus^10^ or severe white matter lesions^11^ was previously shown, and the retrograde transmission of venous pressure waves due to JVR was suggested involved in blood brain barrier disruption^2^. Cerebral venous outflow pathways depend on posture; in the spine position, the IJVs are the primary venous drains; however, in the sitting and standing positions, jugular veins collapse due to high outflow resistance, and the vertebral veins become the major exit pathways^12^. Although JVR has been examined mostly under the Valsalva maneuver in the supine position, spontaneous JVR under normal breathing, as seen during sleep, has a more sustained retrograde-transmitted venous pressure into the cerebral venous system, and may have a greater impact on cerebral circulation and amyloid clearance in daily life. Here, we aimed to clarify whether JVR during normal breathing in the supine position is related to cognitive function and incident dementia in non-demented individuals with vascular risk factors.

## Method

### Study design and patients

Data were derived from a prospective study, the Tokyo Women’s Medical University Cerebral Vessel Disease (TWMU CVD) Registry, from October 2015 to July 2019 (Registration-URL: https://www.umin.ac.jp/ctr/index.htm; UMIN000026671). Written informed consent was obtained from all participants. The study was approved by the Institutional Review Board of Tokyo Women’s Medical University. The research protocol and inclusion criteria of the TWMU CVD registry were previously described in detail^13^. Briefly, we consecutively included patients aged 40 years and older, who presented with cerebral vessel disease on magnetic resonance imaging (MRI) and with one or more cerebrovascular risk factors, such as arterial hypertension, diabetes mellitus, dyslipidemia, coronary artery disease, atrial fibrillation, or smoking. The exclusion criteria for the registry were any kind of aphasia, evidence of dementia (Clinical Dementia Rating^14^ ≥1), and dependence on activities of daily living and walking. We excluded patients who experienced vascular events within 1 month of enrollment. The protocol conformed to the ethical guidelines of the 1975 Declaration of Helsinki in line with the Ethical Guidelines for Epidemiological Research by the Japanese government and the Strengthening the Reporting of Observational Studies in Epidemiology (STROBE) guidelines.

### Cognitive examination

Patients underwent both the Mini-Mental State Examination (MMSE)^15^ and the Japanese version of the Montreal Cognitive Assessment (MoCA-J)^16^ within 3 months of the MRI examination. Among items in MMSE and MoCA-J, we extracted executive function (0-4), memory (0-8), orientation (0-10), language (0-13), visuospatial function (0-5) and attention and working memory (0-11) (**Supplementary Table I**).

### MRI Assessment

Each subject underwent a brain MRI scan within 1 year of entry to the registry. The MRI assessment included white matter hyperintensities (WMH) consisting of periventricular hyperintensities (PVH) and deep white matter hyperintensities (DWMH), lacunar infarctions (LI), cerebral microbleeds (CMBs), and medial temporal atrophy (MTA). The degree of WMH severity was visually rated using axial fluid-attenuated inversion recovery (FLAIR) images. Based on the Fazekas scale (0 = none; 1 = mild; 2 = moderate; 3 = severe)^17^, PVH and DWMH were scored as 0-3; the total WMH score was 0-6 with the sum of PVH and DWMH. The degrees of WMH in the frontal, temporal, parietal, and occipital lobes were visually rated using the Scheltens Scale^18^ with slight modifications: scores of 0-6 (0 = absent, 1 = <3 mm in ≤5, 2 = <3 mm in ≥6, 3 = 4–10 mm in ≤5, 4 = 4–10 mm in ≥6, 5 = ≥11 mm in ≥1, and 6 = confluent) were assigned for deep WMH (frontal, temporal, parietal, and occipital). Lesions in the basal ganglia, internal capsule, centrum semiovale, or brainstem, with hypointensity on T1-weighted imaging (T1WI), hyperintensity on T2-weighted imaging (T2WI), and a hyperintense rim around the cavity on FLAIR were defined as LI^19^; the sizes of which were from 3-15 mm. CMBs were defined as punctate or small patchy lesions <10 mm in diameter with clearly low intensity on T2*WI^19^. MTA was visually evaluated as described by Kim et al.^20^ All MRI feature grades were rated by two trained board-certified neurologists (M.H. and M.S.), who were blinded to the clinical details. The interrater κ for each CSVD feature or MTA score was between 0.80-0.85. A third rater (K.K.) was consulted in cases of disagreement.

### Carotid Duplex sonography

Color-coded duplex sonography was performed using a 7.5-MHz linear transducer (Hitachi Aloka Prosound α7; Hitachi-Aloka Medical, Ltd. Tokyo, Japan). On examination, the subjects were placed in a head-straight, flat supine position. After routine examination of both carotid arteries for the measurement of intima-media complex thickness, % stenosis (if present), and peak systolic blood flow of the common, internal carotid, and vertebral arteries, bilateral IJVs were examined 5 cm above the junction of the IJV and brachiocephalic vein, as described previously with slight modification^8)^. The cross-sectional lumen was recorded, and the probe was turned by 90^0^ at the same site, the IJV was insonated longitudinally, and the color box adjusted to include the entire IJV lumen. The Doppler spectrum was then measured for at least three cardiac cycles under normal expiration. We divided the flow pattern into four groups (**Figure 1**); anterograde flow during the entire cardiac cycle without retrograde flow (Grade A; **Figure 1A**), anterograde flow during the entire cardiac cycle with intermittent retrograde flow (Grade B; **Figure 1B**), a to-and-flow pattern, in which anterograde and retrograde flow appeared alternatively during the cardiac cycle (Grade C; **Figure 1C**), and retrograde flow during the entire cardiac cycle without anterograde flow (Grade D; **Figure 1D**). The former two, mainly anterograde flow, were designated JVR (-), while the latter two were designated JVR (+). For evaluation of IJV in each patient, the higher JVR grade among both sides was used.

**Figure 1.**
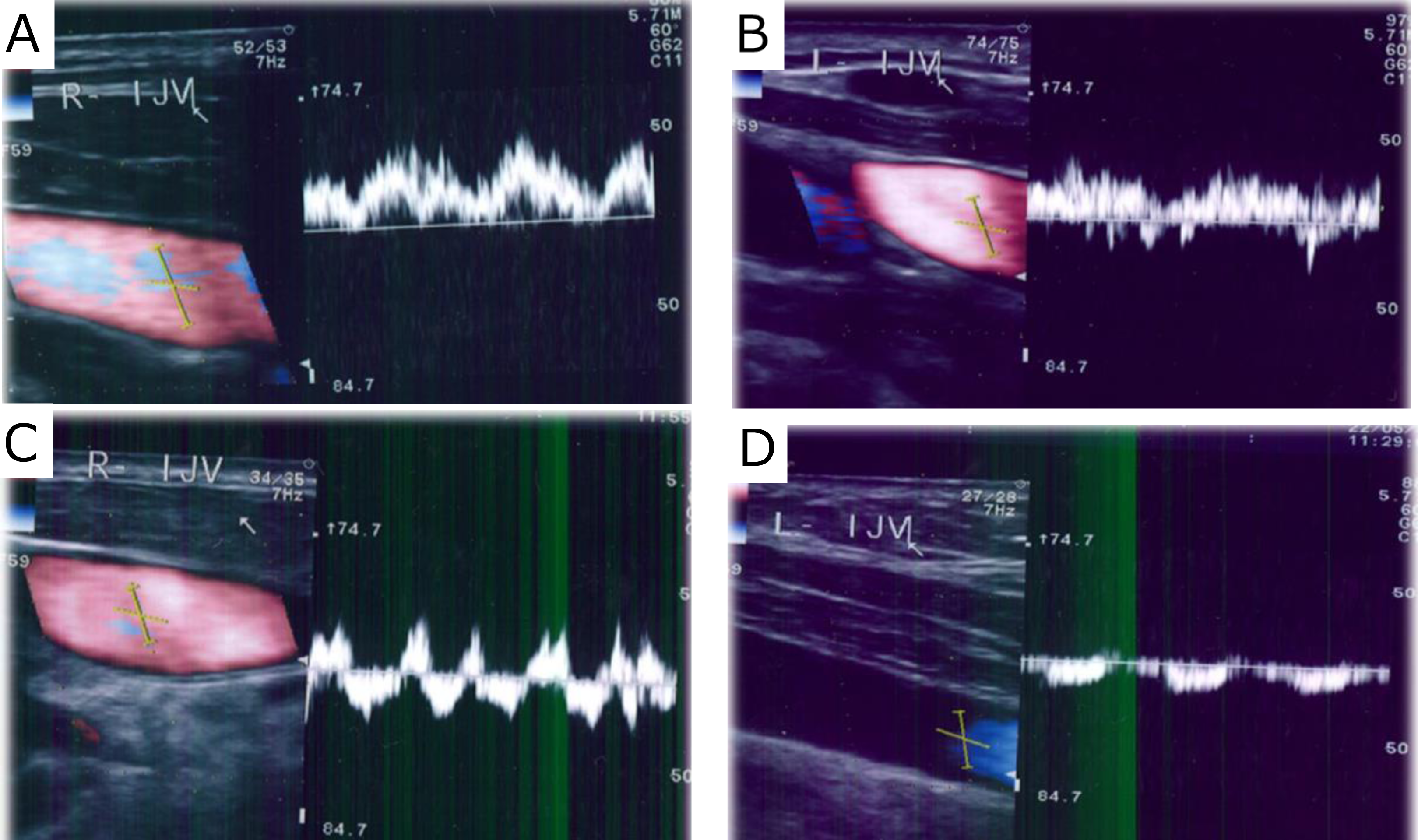
Pattern of internal jugular vein blood flow. (A) Anterograde flow during the entire cardiac cycle without retrograde flow, (B) Anterograde flow during the entire cardiac cycle with intermittent retrograde flow, (C) To-and-flow pattern in which anterograde and retrograde flow appear alternatively during the cardiac cycle, (D) Retrograde flow during the entire cardiac cycle without anterograde flow. The former two, mainly anterograde flow, are designated jugular vein reflux (JVR) (-), while the latter two are designated JVR (+).

An interrater reproducibility test of IJV in 40 vessels of 20 patients (two measurements performed within 30 min) by two blinded examiner (U.A. and S.T.) showed an excellent reproducibility; concordance rates among 4 grades and between JVR (-) and (+) groups, were 100% (40/40), and 97.5% (39/40), respectively. An intrarater reproducibility test (U.A.) of IJV in 128 vessels of 64 patients (two measurements performed blindly at intervals between 6 months and 1 year) also showed a fair reproducibility (concordance rate among four grades and between JVR (-) and (+) groups were 81.3% (104/128) and 97.7%, (125/128), respectively.

### Diagnosis of dementia

Cognitive status was assessed prospectively by a neurologist using the MMSE and CDR^14^. Patients visited outpatient clinic settings to control risk factors every 3 months for prevention of stroke. Changes in patients’ general medical condition were obtained yearly through medical records and interview. Furthermore, several aspects of everyday cognitively driven functioning to rate patients on the CDR were assessed at every clinical visit. Thus, annual evaluations were performed by trained neurologists and included a medical history, CDR score determination, and neurological examination. Patients were cognitively screened with the MMSE at follow-up visits. The final follow-up data were collected on March 2022. During the follow-up periods, patients with suspected cognitive decline were periodically examined by a neurologist. Clinically significant cognitive impairment was defined as MMSE score < 24 or a decline ≥4 points. In addition, patients were considered to have probable dementia if they had 2 consecutive semiannual CDR scores ≥1 and did not revert back to normal cognition. To avoid missing incident dementia cases, we also continuously monitored the medical records of all participants at our clinic or other clinic to obtain information on diagnosed dementia and stroke or death. Furthermore, for those patients who could not attend the clinical visit, a phone interview collecting clinical data was performed with the patient and the carer, whenever possible. Finally, an independent committee of neurologists reviewed all potential cases of dementia with all available information to reach a consensus on the diagnosis, according to the DSM-5^21^. Dementia subtypes were diagnosed according to standardized criteria^22,23^. Time to dementia was defined as the time between the baseline visit and the date of dementia diagnosis. In addition, patients were followed until death or refusal of further participation. Those who did not progress to dementia were censored at their last visit.

### Statistical analysis

All analyses were performed using JMP 14 Pro. For baseline characteristics, continuous variables are expressed as mean and standard deviation (SD), while categorical variables are expressed as frequency and percentage. Ordinal and non-normally distributed variables are shown as median and interquartile range. Relationships between the MMSE, MoCA-J, or each cognitive domain and continuous variables were examined using Person‘s correlation analysis. For categorical variables, differences in MMSE and MoCA-J scores were examined using an unpaired t-test. Furthermore, a linear mixed model was used to examine the associations between JVR and MMSE or MoCA-J scores by controlling for age, sex, education, risk factors, and MRI findings. The Kaplan-Meier method with log-rank tests was used to compare dementia-free survival for JVR.

## Results

A total of 302 patients (71.1±10.5 years; 177 men, 125 women) underwent carotid ultrasonography with IJV evaluations (**Supplementary Figure I**). Their demographic and clinical characteristics are presented in **Table 1**.

**Table 1:** Baseline characteristics of patients.

| Characteristics | All (n=302) | Jugular Vein Reflex |  | P value |
| --- | --- | --- | --- | --- |
|  |  | (-) (n=211) | (+) (n=91) |  |
| Age, years | 71.1±10.5 | 66.7±10.4 | 71.8±9.9 | <0.001 |
| Sex, men, % | 58.6 | 56.8 | 62.9 | 0.363 |
| Education, years | 14.4±2.5 | 14.4 ±2.6 | 14.7±2.4 | 0.285 |
| Hypertension, % | 69.4 | 69.5 | 69.4 | 0.946 |
| SBP, mmHg | 136.9±18.8 | 135.9±20.8 | 137.9±18.7 | 0.522 |
| DBP, mmHg | 75.7±11.9 | 76.3±12.3 | 74.2±10.9 | 0.126 |
| Diabetes mellitus, % | 26.6 | 25.4 | 29.4 | 0.447 |
| HbA1c, % | 6.3±0.9 | 6.2±0.9 | 6.4±1.0 | 0.304 |
| Dyslipidemia, % | 56.3 | 55.4 | 57.6 | 0.680 |
| LDL-C, mg/dl | 105.2±28.1 | 105.7±27.8 | 106.1±29.0 | 0.730 |
| HDL-C, mg/dl | 63.5±20.1 | 63.5±21.3 | 63.5±16.8 | 0.826 |
| TG, mg/dl | 115.9±59.1 | 116.0±60.6 | 115.7±55.2 | 0.850 |
| Atrial fibrillation, % | 9.1 | 8.5 | 10.7 | 0.751 |
| Chronic kidney disease, % | 49.8 | 48.3 | 53.5 | 0.419 |
| eGFR | 60.7±17.2 | 61.4±17.2 | 59.1±17.3 | 0.461 |
| Cigarette current smoking, % | 6.7 | 8.1 | 3.6 | 0.167 |
| Previous cerebrovascular disease, % | 52.9 | 38.9 | 46.5 | 0.187 |
| Previous ischemic heart disease, % | 13.2 | 10.6 | 19.3 | 0.034 |
| Medication, statin use, % | 61.0 | 60.1 | 64.0 | 0.538 |
| Anti-platelet use, % | 61.6 | 62.0 | 60.5 | 0.804 |
| Anti-coagulant use, % | 11.9 | 11.2 | 14.0 | 0.487 |
| **MRI findings** |  |  |  |  |
| Fazekas WMH, median IQR (0-6) | 2 (2-2) | 2 (2-2) | 2 (2-2) | 0.806 |
| Frontal WMH, median IQR (0-12) | 4 (2-6) | 4 (1-6) | 4 (2-6) | 0.485 |
| Parietal WMH, median IQR (0-12) | 2 (0-3) | 2 (0-2) | 2 (0-4) | 0.070 |
| Temporal WMH, median IQR (0-12) | 0 (0-2) | 0 (0-2) | 0 (0-2) | 0.381 |
| Occipital WMH, median IQR (0-12) | 0 (0-2) | 0 (0-2) | 1 (0-2) | 0.005 |
| Presence of lacunae, % | 45.8 | 43.3 | 51.7 | 0.184 |
| Presence of microbleeds, % | 29.2 | 28.0 | 31.9 | 0.535 |
| Temporal lobe atrophy,<br>median IQR(0-6) | 2 (1-4) | 2(0-4) | 2 (2-3) | 0.609 |
| Presence of major cerebral<br>artery lesion, % | 12.9 | 14.4 | 9.3 | 0.235 |

Abbreviations: SBP = systolic blood pressure; DBP = diastolic blood pressure; LDL-C = low-density lipoprotein cholesterol; HDL-C = high-density lipoprotein cholesterol; TG = triglyceride; eGFR = an estimated glomerular filtration rate; WMH = white matter hyperintensity.
\*Chronic kidney disease was defined as eGFR of less than 60 ml per minute per 1.73 m<sup>2</sup> of body-surface area.

Among the 302 patients, 122 (40.4 %) showed Grade A in IJV, 89 (29.5%) Grade B, 76 (25.2%) Grade C, 15 (5.0%) Grade D (Figure 1A-D). In total, 91 (30.1%) showed JVR (Figure 1C, 1D) on either side. The jugular flow patterns of the 604 vessels in 302 patients are shown in **Supplementary Table II**. JVR was found more frequently on the left side (20.9%, 63/302) than on the right side (13.2%, 42/302). Fourteen patients (4.6%) showed bilateral JVR, while 77 (25.5%) showed a unilateral JVR. The incidence of JVR increased with age (**Figure 2A**). Patients with JVR were older (P<0.001), and more likely had a history of previous ischemic heart disease (P=0.034) than those without JVR. (**Table 1**). In terms of MRI findings, the Fazekas WMH scores were similar between the two groups, but the occipital WMH grades were more severe in patients with versus without JVR (P=0.005). (**Table 1**).

**Figure 2:**
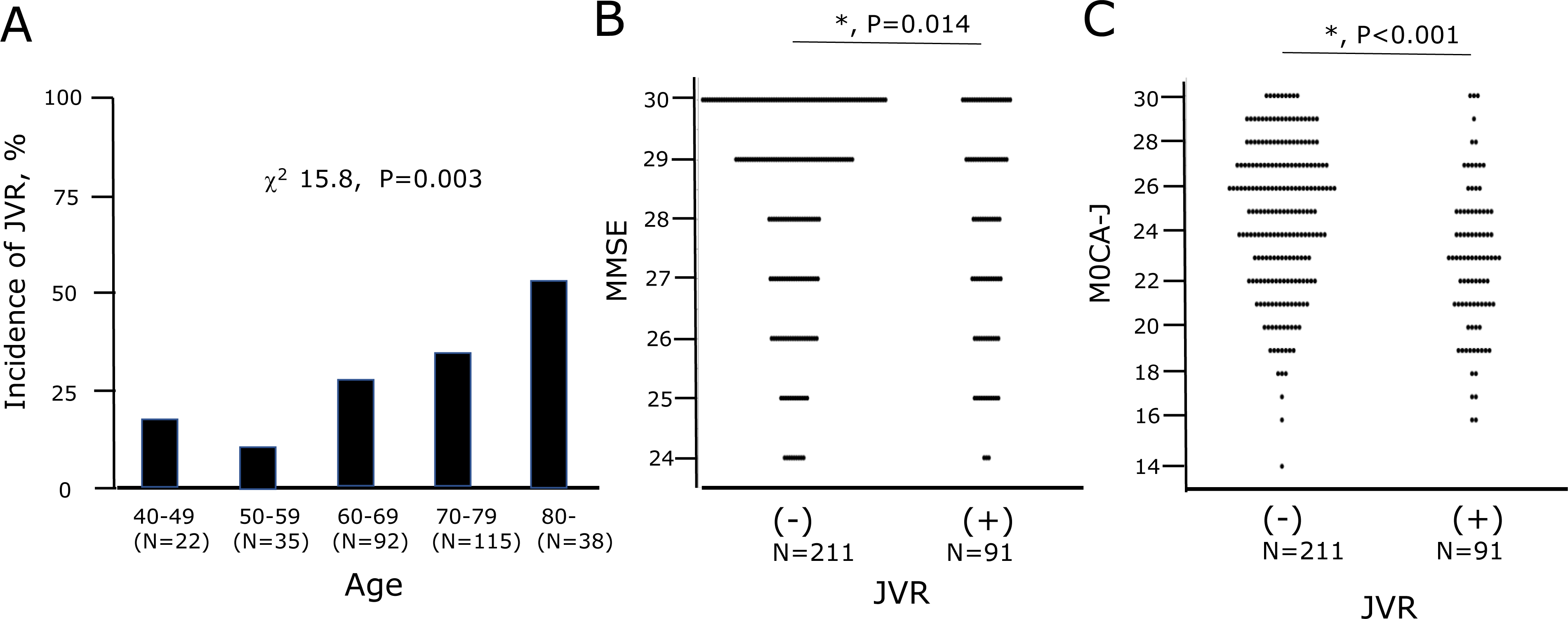
(A) Incidence of jugular vein reflux (JVR) by age, (B) Mini-mental State Examination (MMSE) score and (C) Montreal Cognitive Assessment-Japanese (MoCA-J) score in patient groups with versus without JVR.

MMSE scores were 28.5±1.7, 28.2±1.9, 27.9±1.8 and 27.0±2.0 in Grade A, B, C and D, respectively (ANOVA, P=0.107). MoCA-J scores were 25.2±3.1, 24.3±3.3, 23.1±3.2 and 21.6±2.5 in Grade A, B, C and D, respectively (ANOVA, P<0.001). Global cognitive function and each subdomain in JVR (-) and JVR (+) groups are shown in **Table 2**. The distribution patterns of MMSE and MoCA-J scores between the JVR (-) and JVR (+) groups are shown in **Figures 2B and 2C**. Mean MMSE and MoCA-J scores were significantly lower in the JVR (+) versus JVR (-) group, while the mean executive function and memory scores were significantly lower in the JVR (+) versus JVR (-) group. The scores of the other subdomains were similar between groups.

**Table 2:**
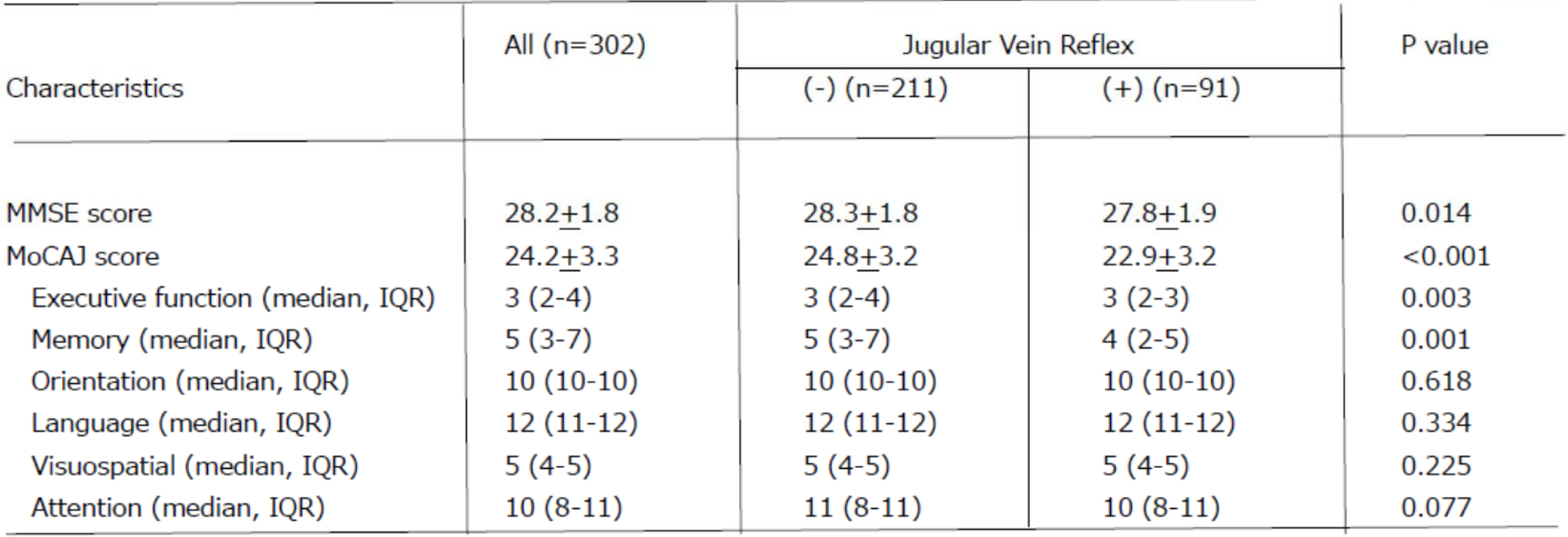
Cognitive function of patients.

| Characteristics | All (n=302) | Jugular Vein Reflex |  | P value |
| --- | --- | --- | --- | --- |
|  |  | (-) (n=211) | (+) (n=91) |  |
| MMSE score | 28.2±1.8 | 28.3±1.8 | 27.8±1.9 | 0.014 |
| MoCAJ score | 24.2±3.3 | 24.8±3.2 | 22.9±3.2 | <0.001 |
| Executive function (median, IQR) | 3 (2-4) | 3 (2-4) | 3 (2-3) | 0.003 |
| Memory (median, IQR) | 5 (3-7) | 5 (3-7) | 4 (2-5) | 0.001 |
| Orientation (median, IQR) | 10 (10-10) | 10 (10-10) | 10 (10-10) | 0.618 |
| Language (median, IQR) | 12 (11-12) | 12 (11-12) | 12 (11-12) | 0.334 |
| Visuospatial (median, IQR) | 5 (4-5) | 5 (4-5) | 5 (4-5) | 0.225 |
| Attention (median, IQR) | 10 (8-11) | 11 (8-11) | 10 (8-11) | 0.077 |

The MMSE and MoCA-J scores with confounding factors are shown in **Supplementary Table III**. Age was significantly associated with both MMSE and MoCA-J scores; mean MMSE scores were higher in patients with high education, while mean MoCA-J scores were higher in women. Other vascular risk factors did not show a significant relationship with MMSE and MoCA-J scores among the 302 patients.

The association between JVR and MoCA-J score remained significant after the adjustment for confounding factors (**Table 3**). The association between JVR and MMSE was significant after the adjustment for age and sex; however, the association was no longer significant after the further adjustment for education and risk factors (**Table 3)**.

**Table 3.**
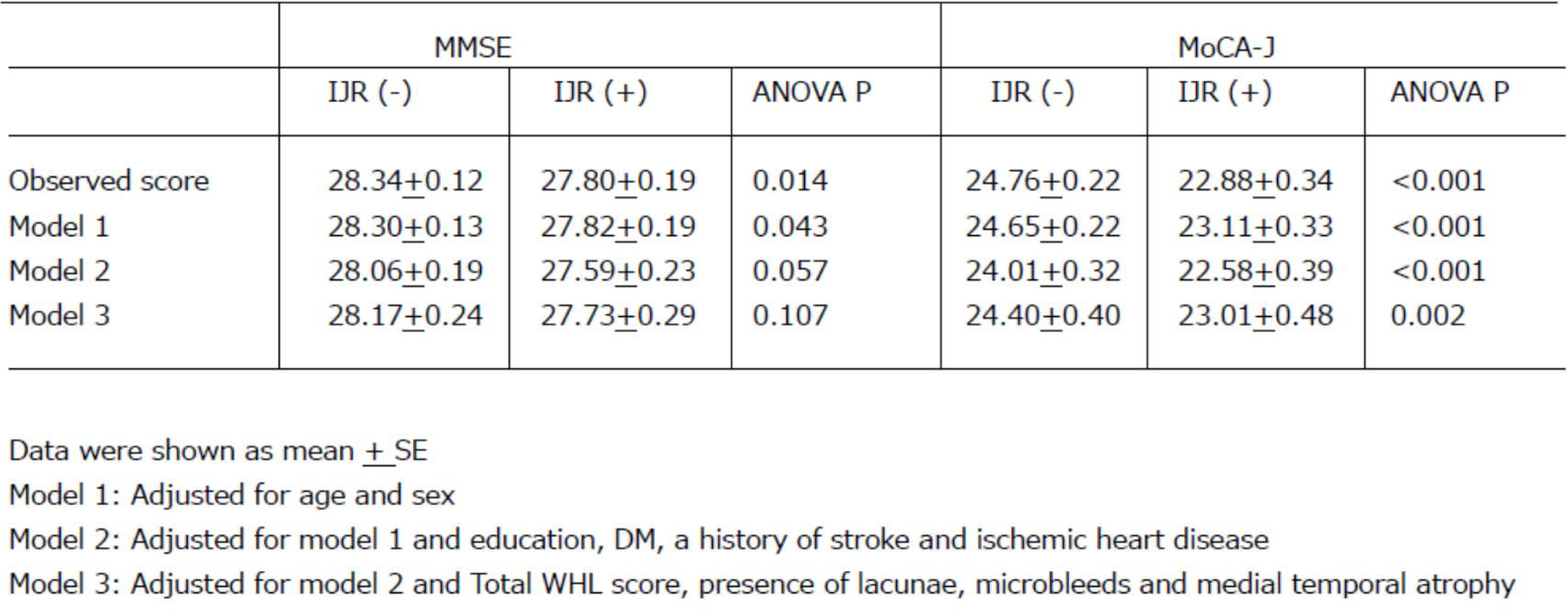
Associations of MMSE and MOCA-J with jugular vein reflux.

From the initial cohort (N=302), 13 patients (4.3%) withdrew consent, five patients (1.7%) were lost, and four patients (1.3%) had died during follow-up. 81.1% (n=245) of the patients could be examined in a clinical visit at the end of follow-up. Among these 245 patients, 25 patients had a decline in MMSE score of ≥4 points, or had an MMSE score <24 by the end of follow-up. These patients were subsequently examined for CDR score determination, and 9 were diagnosed with dementia. Also using data from telephone interview or medical records, we ascertained cognitive status in the remaining 35 patients (11.6% of initial sample), and 2 patients were diagnosed with dementia. Thus, during the median 5.2-year follow-up, all-cause dementia was diagnosed in 11 patients (Alzheimer disease: 10, vascular dementia: 1). The survival analyses for dementia-free rate curves created with respect to JVR are shown in Figure 3. Although no significant difference was seen between JVR (-) and JVR (+) groups (Figure 3A), patients with severe JVR were significantly more likely to be diagnosed with dementia compared with other groups (Figure 3B, log-rank test, P=0.006).

**Figure 3:**
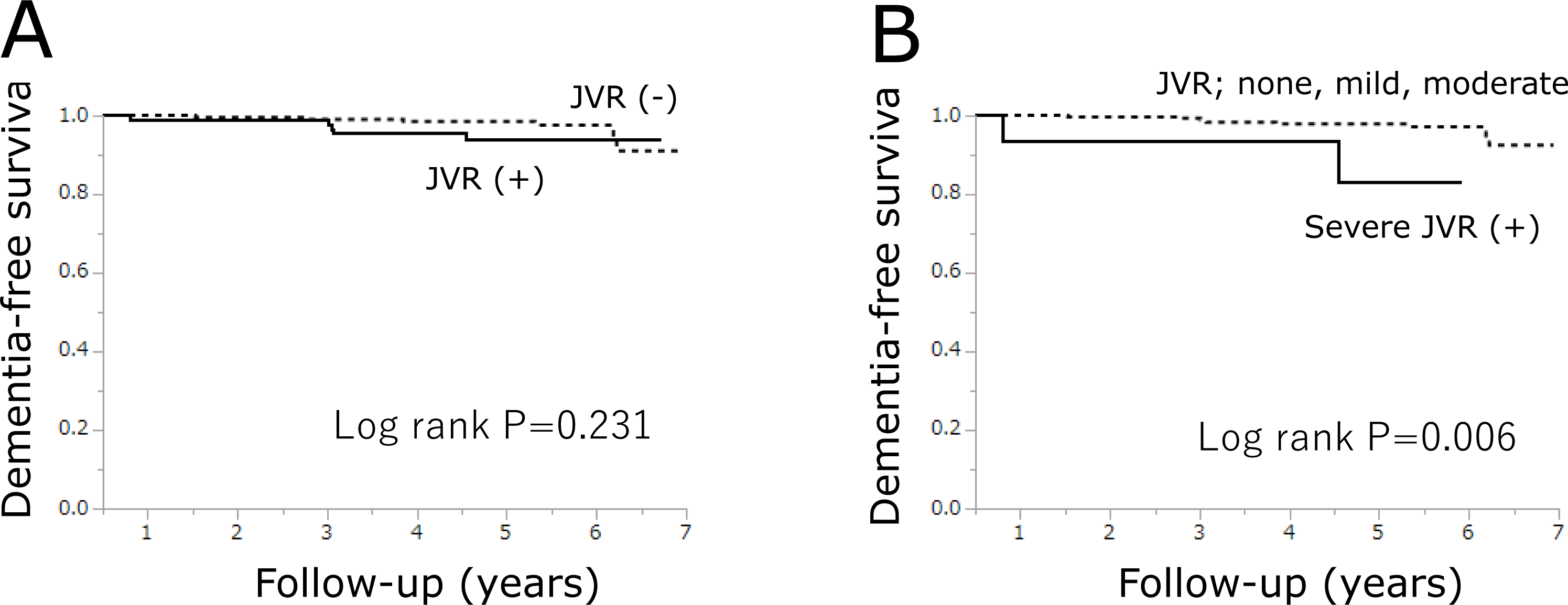
Kaplan-Meier survival analysis of time to dementia diagnosis by baseline jugular vein reflux (JVR) grade.

## Discussion

To the best of our knowledge, this is the first study to clarify the association between spontaneous JVR and cognitive function or incident dementia in individuals without dementia. This study included more than 300 patients; therefore we examined the association after adjusting for confounding factors with JVR and cognitive function.

Several neurological diseases and conditions are associated with cerebral venous insufficiency. Although the correlation between venous insufficiency and multiple sclerosis proposed by Zamboni et al.^24^ was later denied^25^, age-related neurological conditions such as normal hydrocepahus^10^, transient global amnesia^26^, white matter lesions^11^ and cognitive decline in mitral valve regurgitation^27^ were demonstrated associated with JVR.

The incidence of JVR under spontaneous breathing, the Valsalva maneuver, and both in previous studies is summarized in **Supplementary Table IV.** Both age and cardiac dysfunction increase the incidence of JVR^2^. Our study also clearly showed that aging increased the incidence of JVR (Figure 1). Under spontaneous breathing, the incidence of JVR seemed higher on the left versus right side^11,28,29^, whereas under a Valsalva maneuver, it was similar or higher on the right side^8,27,30^. The narrow space between the sternum and the thoracic outlet arteries, especially under normal breathing, might compress the left brachiocephalic vein, leading to JVR in the left internal vein^27^. Full inspiration under the Valsalva maneuver increases the distance between the aortic arch and sternum, thus extinguishing that tendency^31^. In contrast, the right IJV is closer to the heart than the left IJV, and more susceptible to retrograde pressure via the central vein under the Valsalva maneuver, which might explain the higher incidence of JVR in the right IJV in previous studies.

Several mechanisms would underlie the association between the JVR and cognitive function. First, an increase in cerebral venous pressure could decrease the absorption of cerebral spinal fluid through the arachnoid villi as seen in normal pressure hydrocephalus. Second, cerebral venous insufficiency in JVR would influence glymphatic circulation^32^ and impair amyloid β clearance. Third, increased cerebral venous pressure may disrupt the blood-brain barrier, trigger microhemorrhage and decrease cerebral blood flow, resulting in cognitive decline^2^. Our results showed that JVR is related to occipital WMH (Table 1), in line with previous findings.^11^

This study had several limitations. First, our study suggested that severe JVR was associated with incident dementia, but the event number was small and did not allow us to adjust confounding factors such as age and MMSE scores, therefore, further studies will be required to clarify whether JVR predicts incident dementia. Second, the evaluation of white matter lesions and MTA was semi-quantitative. A quantitative evaluation is needed to precisely clarify the association between the JVR and MRI findings. Third, it remains unclear whether JVR intervention recovers cognitive function. And finally, we evaluated the JVR under normal breathing. Therefore, it remains unknown whether JVR during the Valsalva maneuver is associated with cognitive decline. However, in daily life, especially during sleep, we breathe normally, not in full inspiration, in the spine position. The incidence of JVR in the present study reflects the percentage of JVR during sleep in daily life, and would have a greater impact on the cerebral venous circulation than the Valsalva maneuver. In conclusion, here we clarified the relation between JVR with MoCA-J scores independent of age and confounding factors, and suggested the association between severe JVR and incident dementia. Our findings suggest that spontaneous JVR affects cognitive function.

## Data Availability

Data information in this paper is availbale based on the approriate request.

## Nonstandard Abbreviations and Acronyms

IVJ: internal jugular vein
JVR: jugular vein reflux
HDL-C: high-density lipoprotein cholesterol
LDL-C: low-density lipoprotein cholesterol
MRI: magnetic resonance imaging
MMSE: mini-mental State Examination
MoCA-J: Montreal Cognitive Assessment-Japanese
STROBE: Strengthening the Reporting of Observational Studies in Epidemiology
TIA: transient ischemic attack
TWMU: Tokyo Women’s Medical University
CDR: clinical dementia rating

## Acknowledgements

None.

## Supplemental Materials

Table I-IV Figure I

